# Impact of COVID-19 infection on maternal and neonatal outcomes: a review of 287 pregnancies

**DOI:** 10.1101/2020.05.09.20096842

**Authors:** Fatemeh Azarkish, Roksana Janghorban

**Author notes:** **Corresponding Author: Roksana Janghorban** PhD., Associate Professor in Reproductive Health, Maternal-Fetal Medicine Research Center, Midwifery Department, School of Nursing and Midwifery, Nemazee Square, Zand Blv., Shiraz University of Medical Sciences, Shiraz, Iran, Facsimile number: 98-7136474252.

## Abstract

Pregnant women are vulnerable group in viral outbreaks especially in the severe acute respiratory syndrome coronavirus 2 (SARS-CoV-2) pandemic. The aim of this review was to identify maternal and neonatal outcomes in available articles on pregnancies affected by COVID-19. The articles that had assessed outcomes of pregnancy and perinatal of women with COVID-19 between Oct 2019 and Apr 30, 2020 without language limitation were considered. All kinds of studies such as case report, case series, retrospective cohort, case control were included. We searched databases, selected relevant studies and extracted data regarding maternal and neonatal outcomes from each article. Data of 287 pregnant women with COVID-19 of 6 countries were assessed from 28 articles between December 8, 2019 and April 6, 2020. Most pregnant women reported in their third trimester, 102 (35.5%) cases were symptomatic at the time of admission. Common onset symptoms, abnormal laboratory findings, and chest computed tomography pattern were fever (51.5%), lymphocytopenia (67.9%), and multiple ground-glass opacities (78.5%) respectively. 93% of all deliveries were done through cesarean section. No maternal mortality and 3 % ICU admission were reported. Vertical transmission was not reported but its possibility was suggested in three neonates. One neonatal death, one stillbirth, and one abortion were reported. All newborns were not breastfed. This review showed fewer adverse maternal and neonatal outcomes in pregnant women with COVID-19 in comparison with previous coronavirus outbreak infection in pregnancy. Limited data are available regarding possibility of virus transmission in utero, during vaginal childbirth and breastfeeding. Effect of COVID-19 on first and second trimester and ongoing pregnancy outcomes in infected mothers is still questionable.

## 1. Introduction

Outbreak of Coronavirus (COVID-19) as a new respiratory disease has affected over two million individuals throughout the world and the World Health Organization (WHO) has declared the outbreak as a global pandemic on March11, 2020 [1,2]. Understanding of the effect of viral infection during pregnancy is considerably important especially in pandemic due to possible effect on the pregnant woman and the fetus [3]. Studies showed that pregnant mothers were vulnerable group in viral outbreaks of influenza-A, H1N1, the severe acute respiratory syndrome coronavirus (SARS-CoV), the Middle East respiratory syndrome coronavirus (MERS-CoV), Ebola, and Zika virus and had higher risk of mortality and worse maternal and neonatal outcomes such as abortion, still birth, preterm delivery, and birth defects [4-8].

Some conditions increase vulnerability of mothers and fetuses to become infected with outbreak viruses especially respiratory viruses. The first one is maternal physiologic adaptive changes in cardiopulmonary system during pregnancy which leads to increased heart rate, and stroke volume, and reduced pulmonary residual capacity. These changes could increase the risk of hypoxemia [9]. The second issue is maternal immunologic alterations and a shift from Th1 to Th2 immunity that occurs during pregnancy. This shift could decrease the robustness of cell-mediated immunity, alter responses to viral respiratory infections during pregnancy, and increase severity of these infections [10,11]. The third factor is related to characteristics of innate and adaptive immunity in fetus including lower cytolytic function of fetal natural killer (NK) cells compared to adults, lower intensity of antigen-specific antibody response in comparison with adults, immature T cell immunity with suppressed Th1 responses and upregulated Th2 responses, hyporesponsive macrophages, and scarce soluble inflammatory mediators [12-14]. All of these characteristics increase fetal susceptibility to infections. Furthermore, a viral infection of placenta could produce inflammatory cytokines, activate maternal immune system, impair placenta, induce abortion or preterm labor in perinatal period and lead to long-term neurodevelopmental sequelae in adulthood [15-17].

Despite some similarities among COVID-19, SARS and MERS, the 2019 novel coronavirus seems to be less lethal and more easily spread than the other two coronavirus according to available information [18]. This characteristic could raise critical concern as regards widespread transmission in the community and increase infected pregnant women numbers and the possibility of vertical transmission of COVID-19 from mother to the fetus, and adverse outcome on maternal, fetal and neonatal health.

Current knowledge related to effect of COVID-19 infection on maternal and perinatal outcomes are based on information that exists in some scattered case reports and case series which were mostly reported from Wuhan, Hubei Province, China. It seems that better clinical management of the infection in pregnancy needs to have more information of COVID-19 behavior in a large number of infected pregnant cases in different trimesters from various countries. This review was conducted to identify maternal and neonatal outcomes in available articles on pregnancies affected by COVID-19 throughout the world.

## 2. Materials and Methods

### Eligibility criteria

In this review, the articles that had assessed outcomes of pregnancy and perinatal of women with COVID-19 between Oct 2019 and Apr 30, 2020 were considered. All kinds of studies such as case report, case series, retrospective cohort, case control were included. Non-English language publications were also included and data extraction of them was used through Google Translator.

### Literature search and data extraction

We searched PubMed, Scopus, Web of Science (WOS), and MedRxiv using MeSH-compliant keywords including: "2019-nCoV infection", "coronavirus disease 2019", "COVID-19 pandemic", "2019-nCoV disease", "2019 novel coronavirus disease", "COVID19", "2019 novel coronavirus infection", "coronavirus disease-19", "severe acute respiratory syndrome coronavirus 2", "SARS-CoV-2", "pregnancy", "pregnant women", "maternal", and "prenatal care". Search strategy of them is mentioned in Appendix 1. Relevant studies [19-46] were selected based on titles and abstracts, then their full texts were assessed by two reviewers. The following data were extracted from each article: name of the first author, time of study, the number of mothers and neonates, mother’s age, gestational age, delivery mode, maternal and neonatal outcomes, and COVID-19 vertical transmission (Table 1).

**Table 1.**
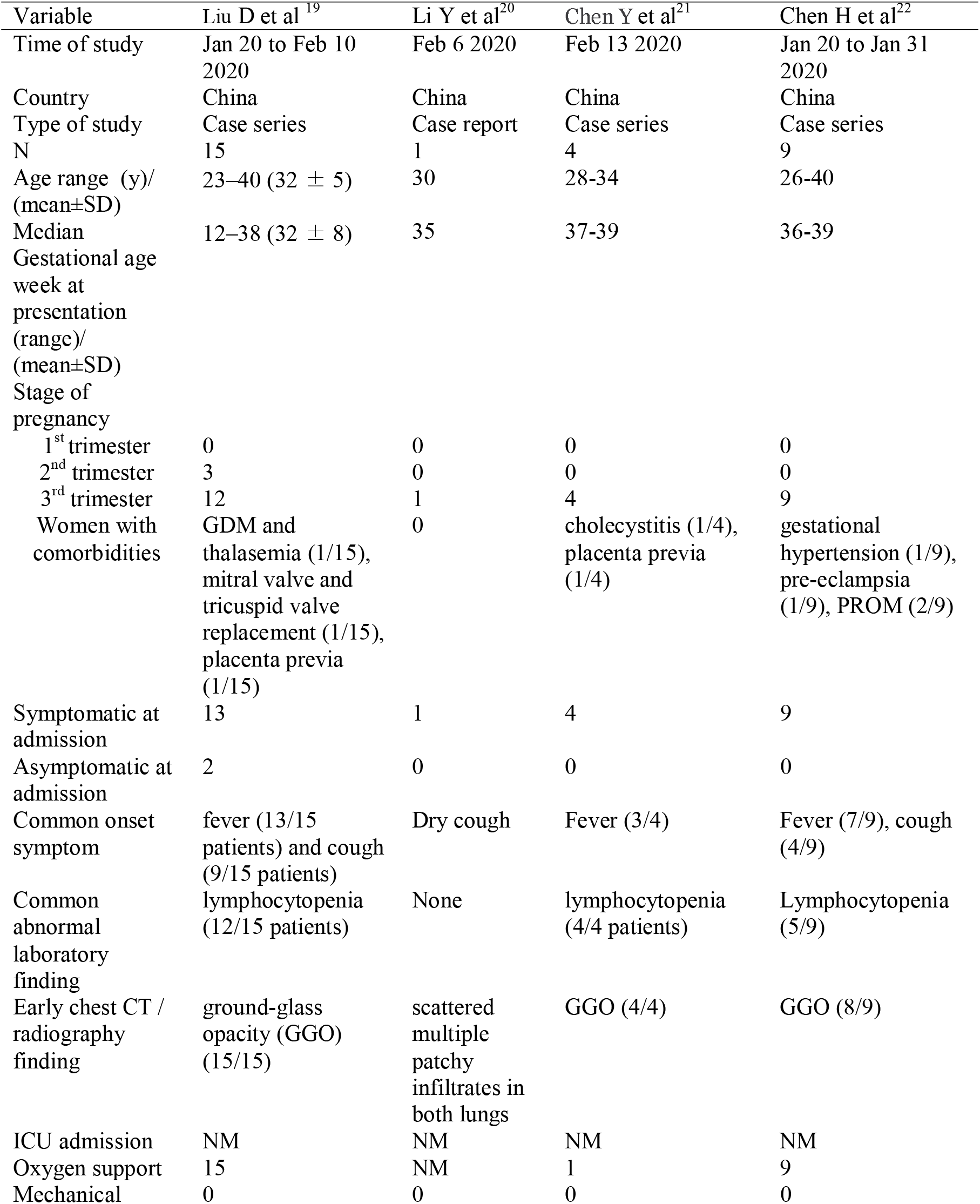
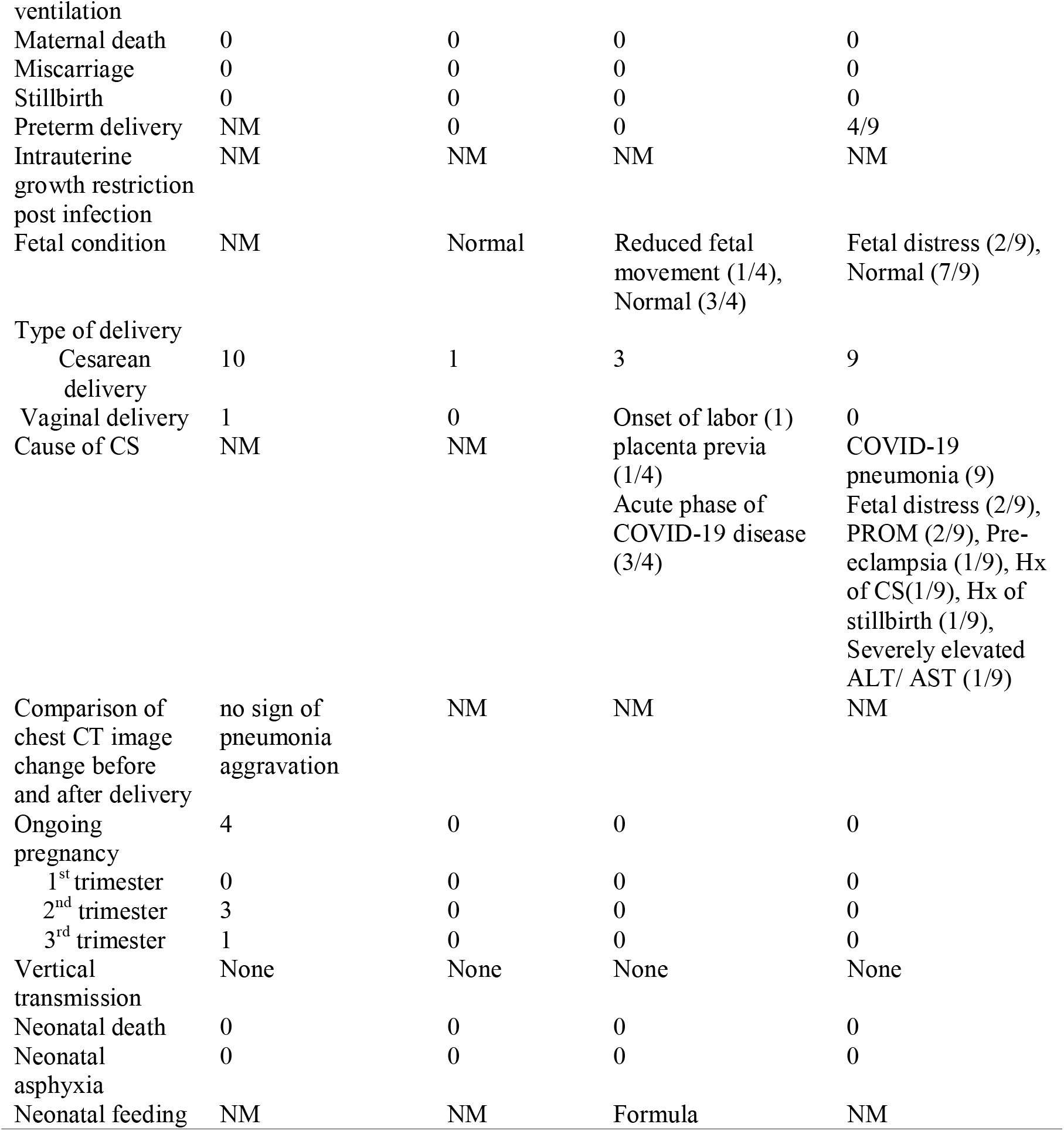
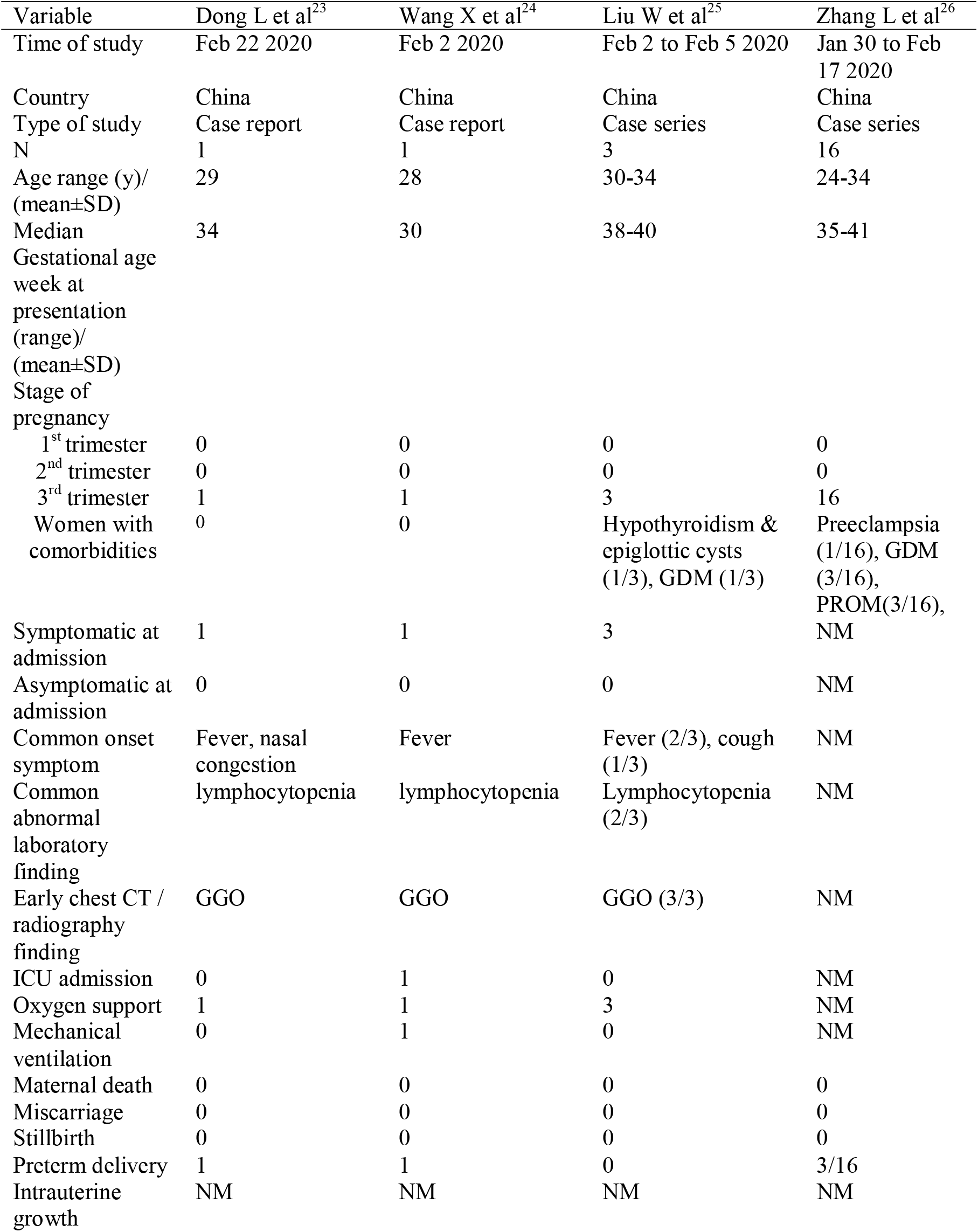
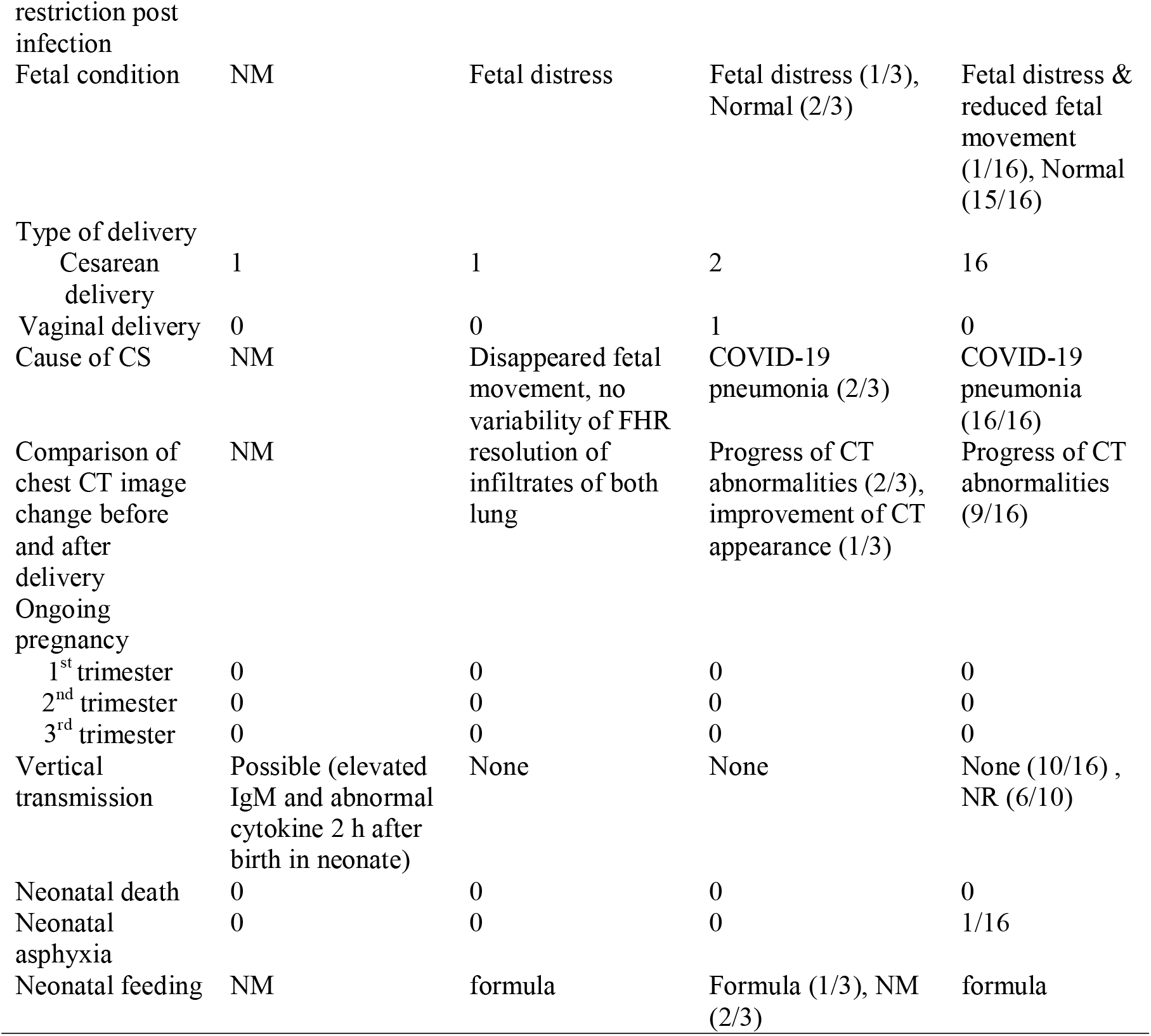
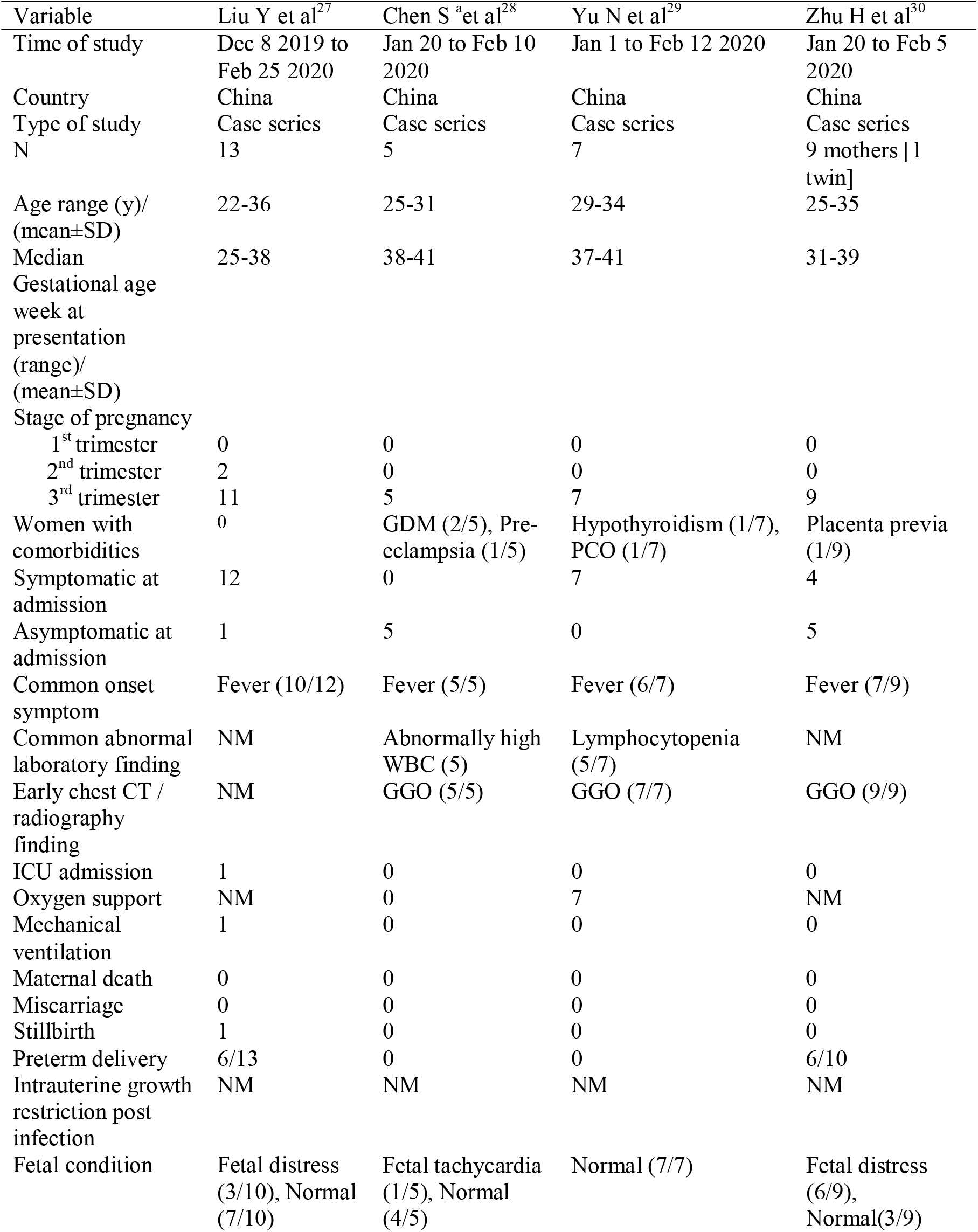
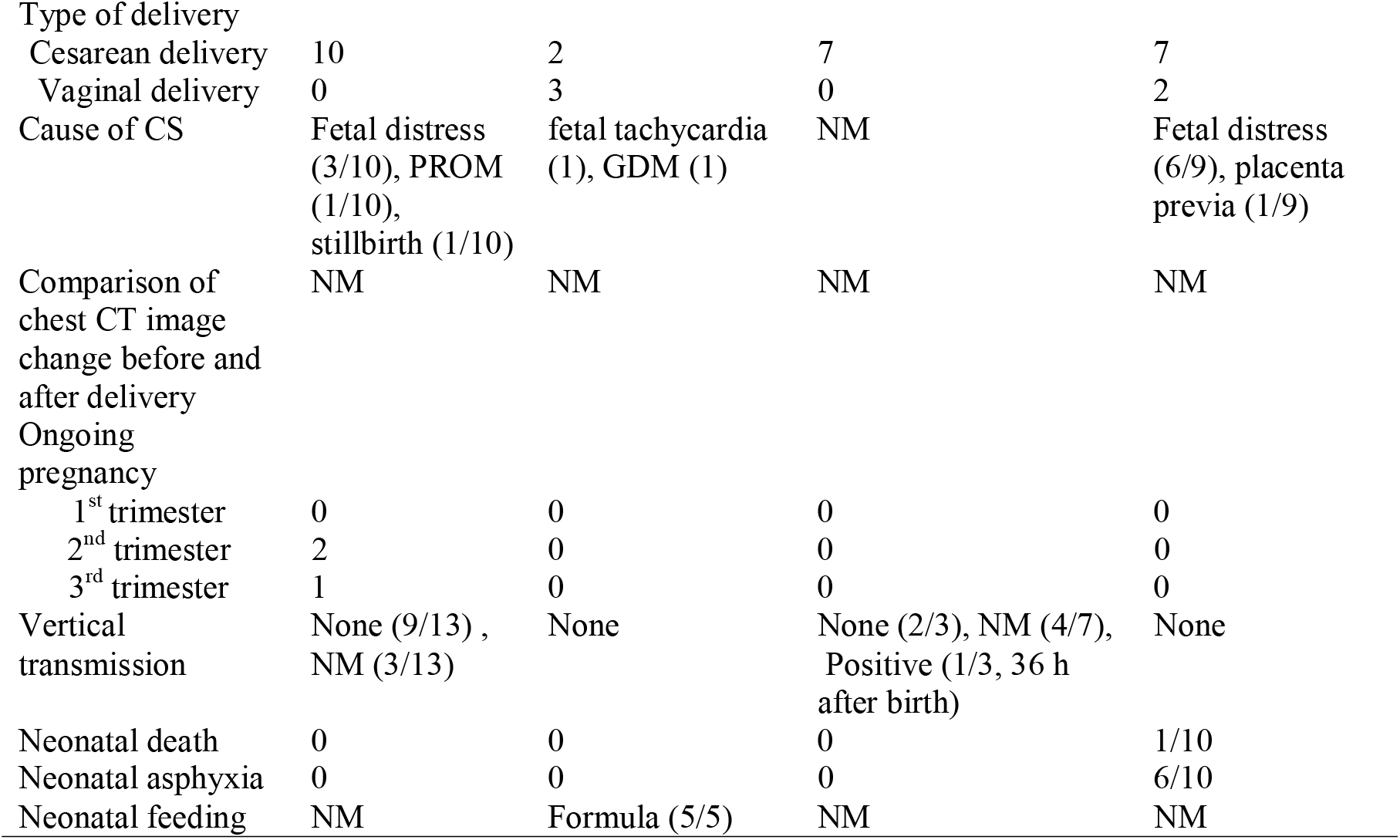
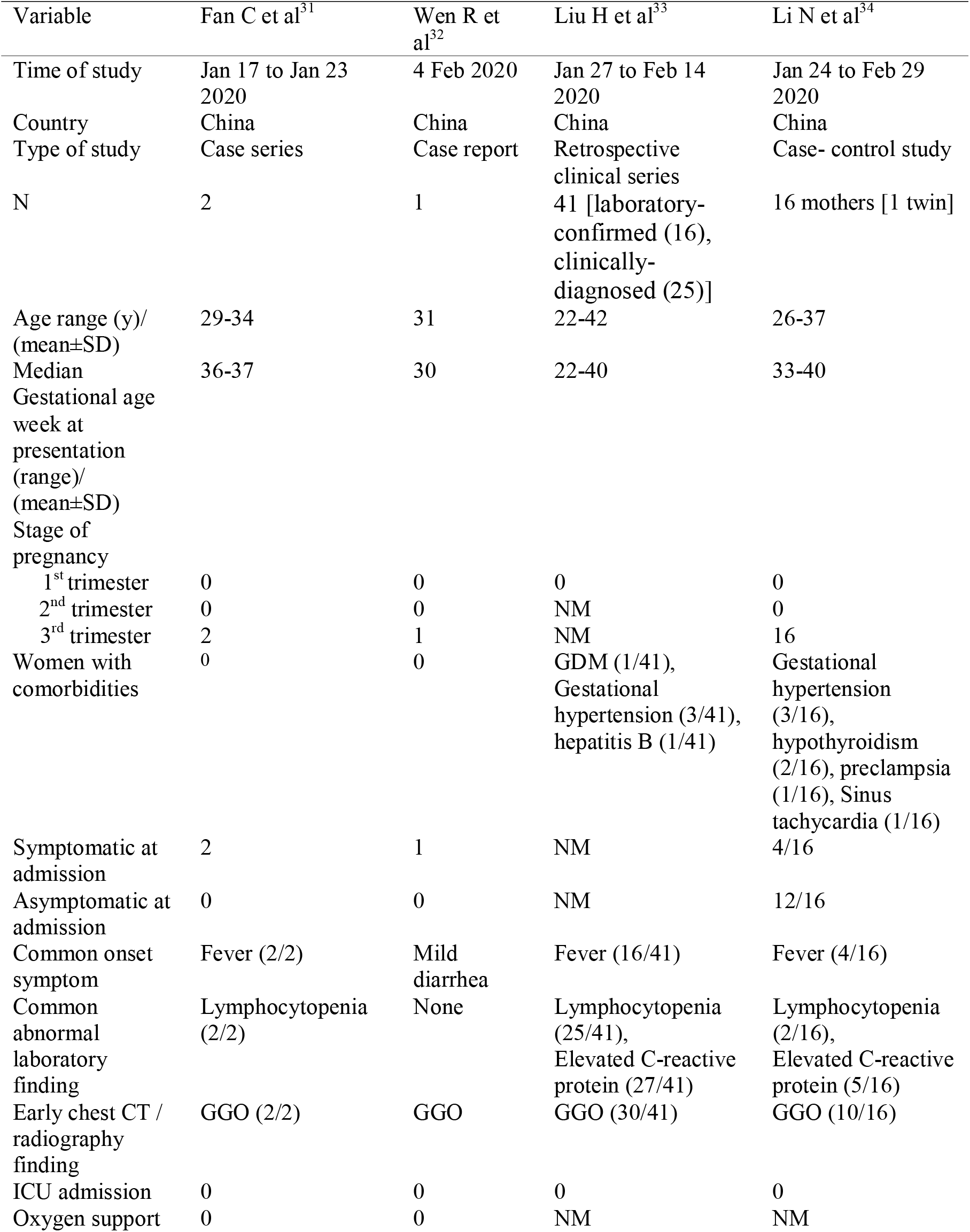
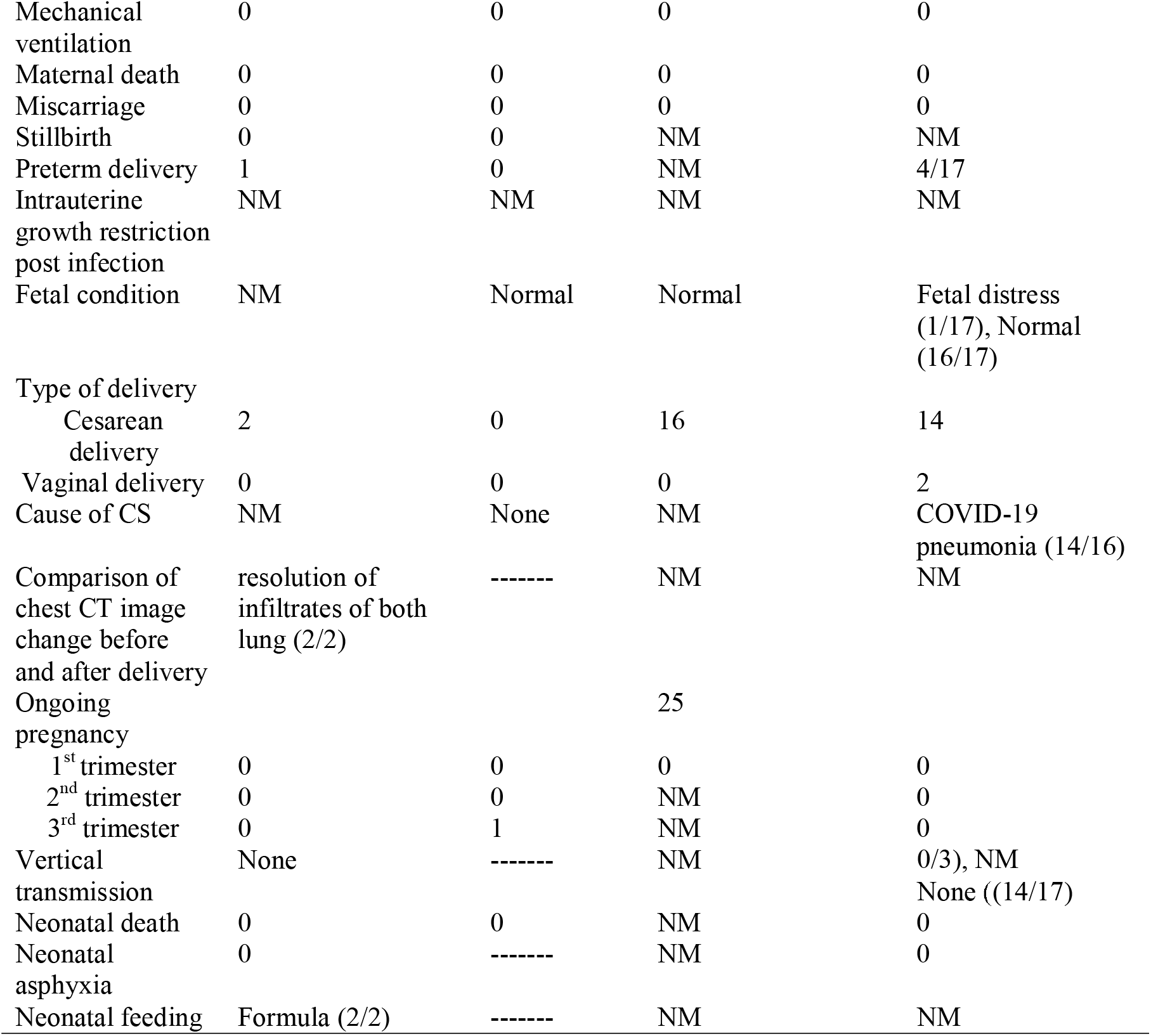
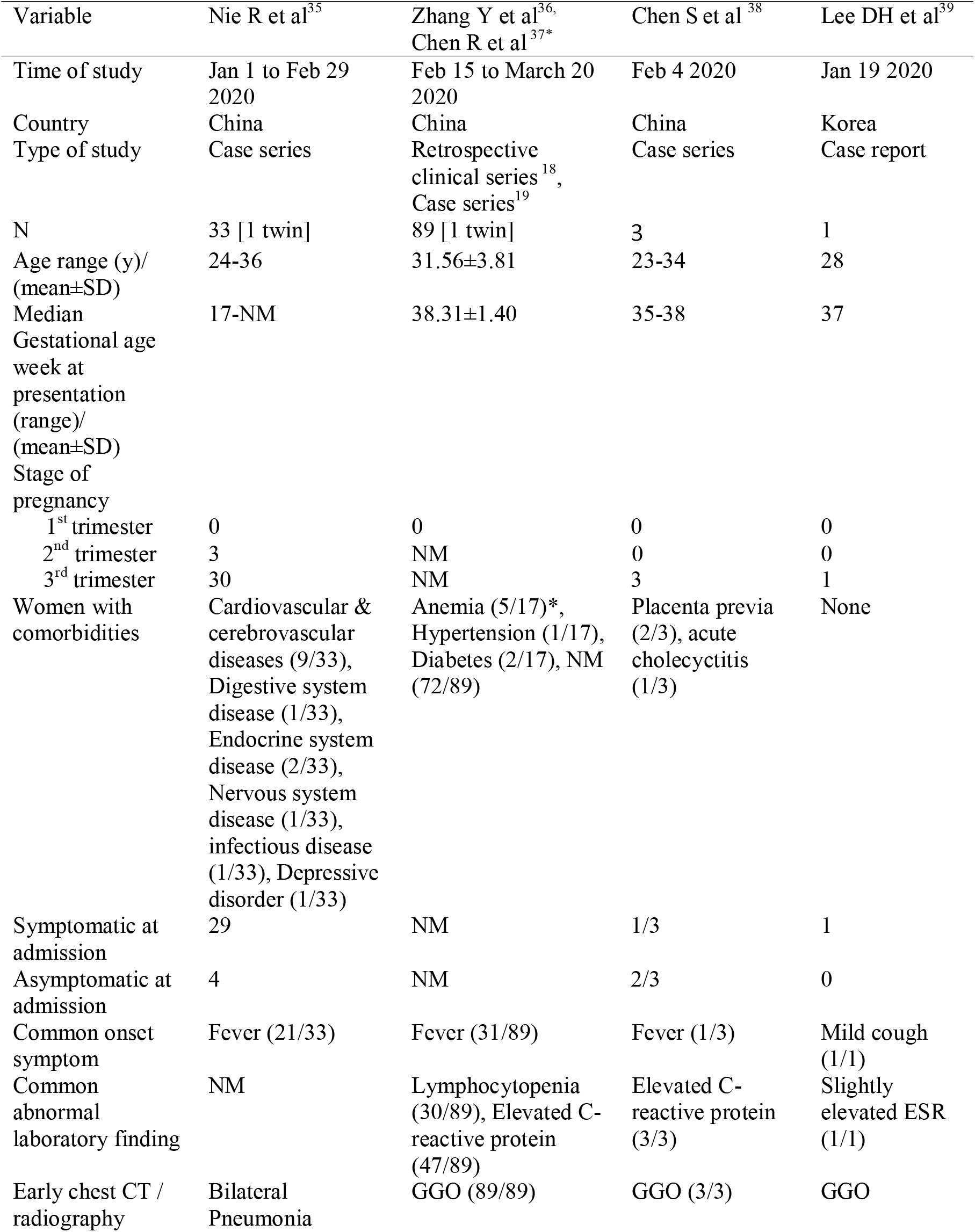
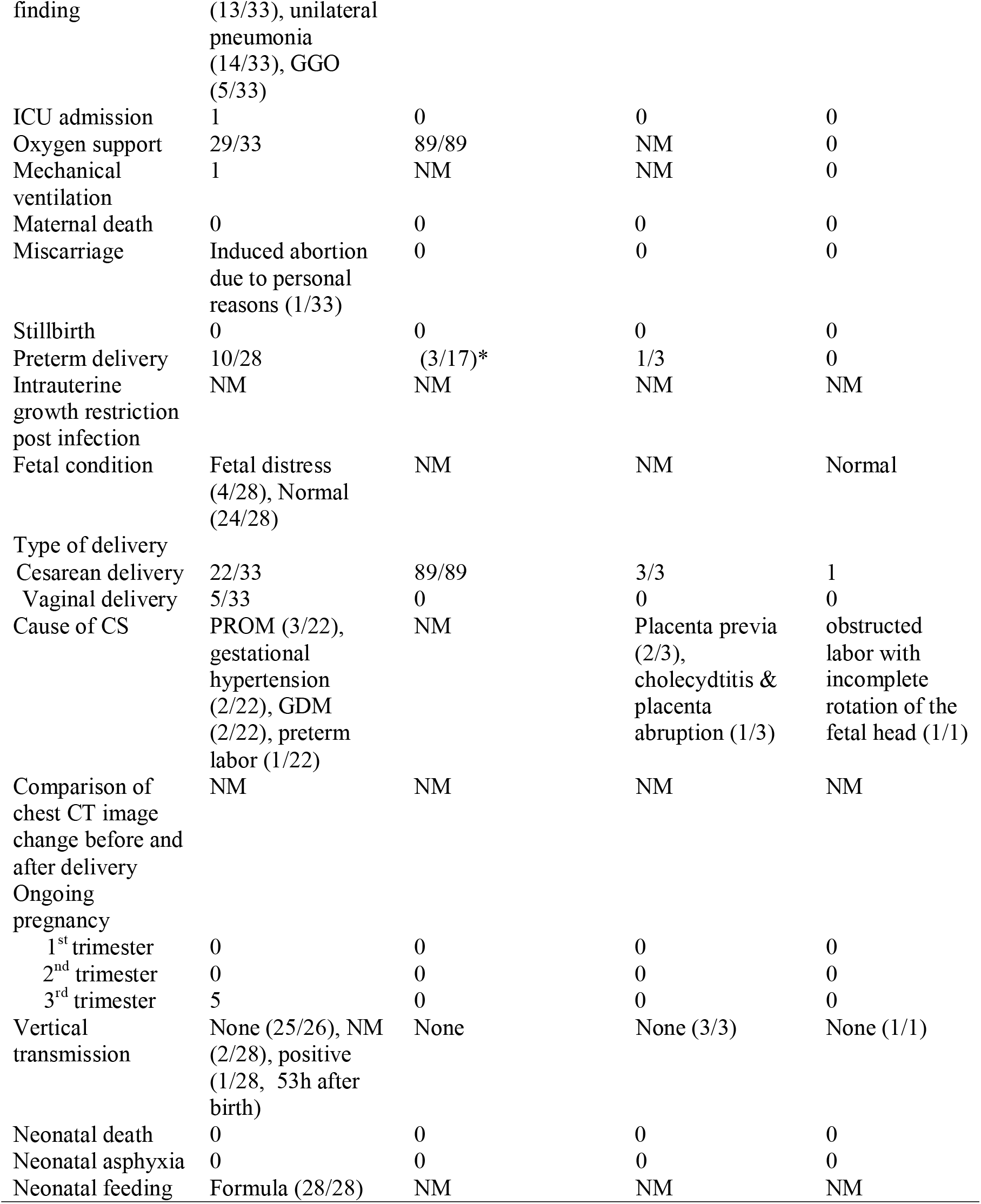
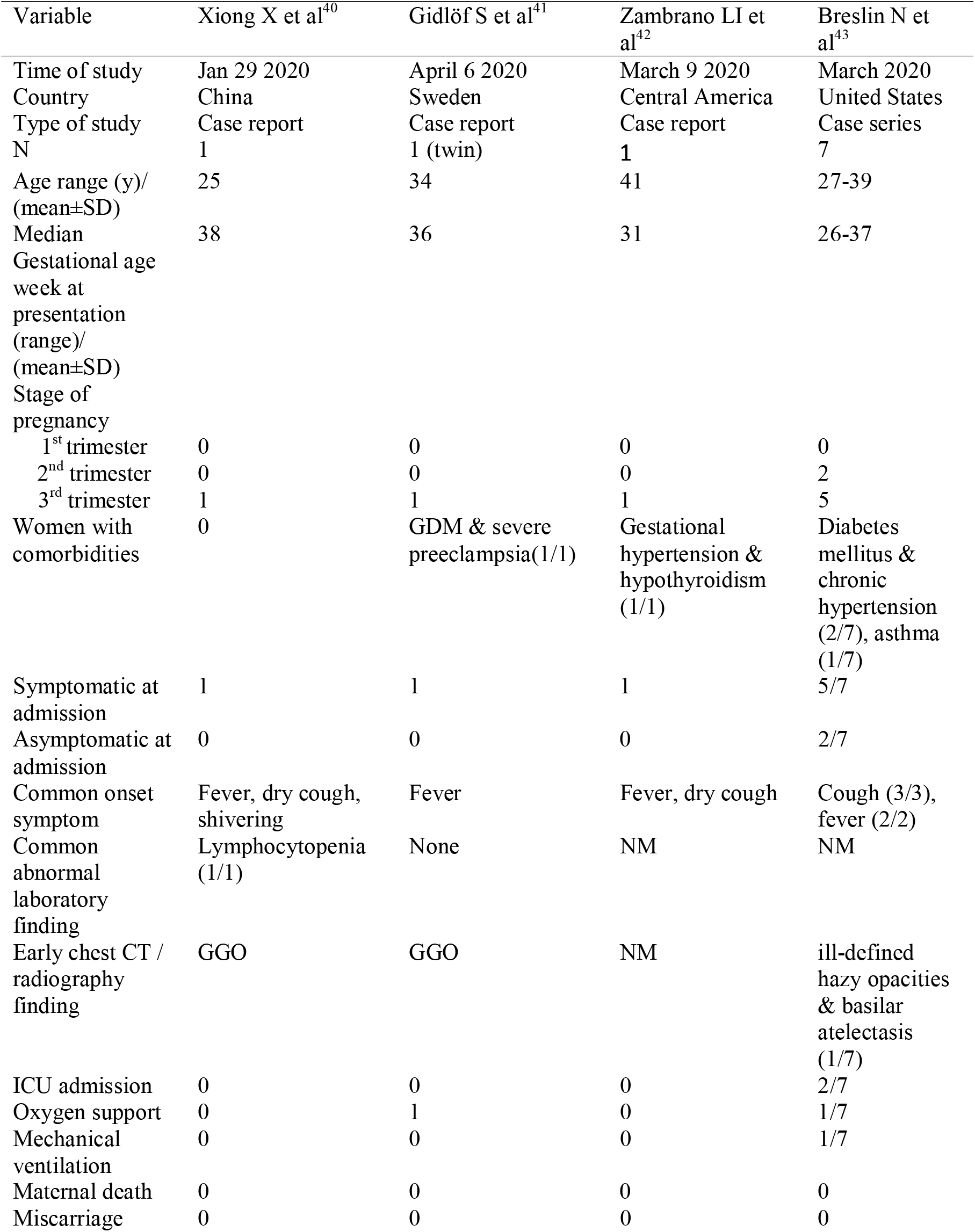
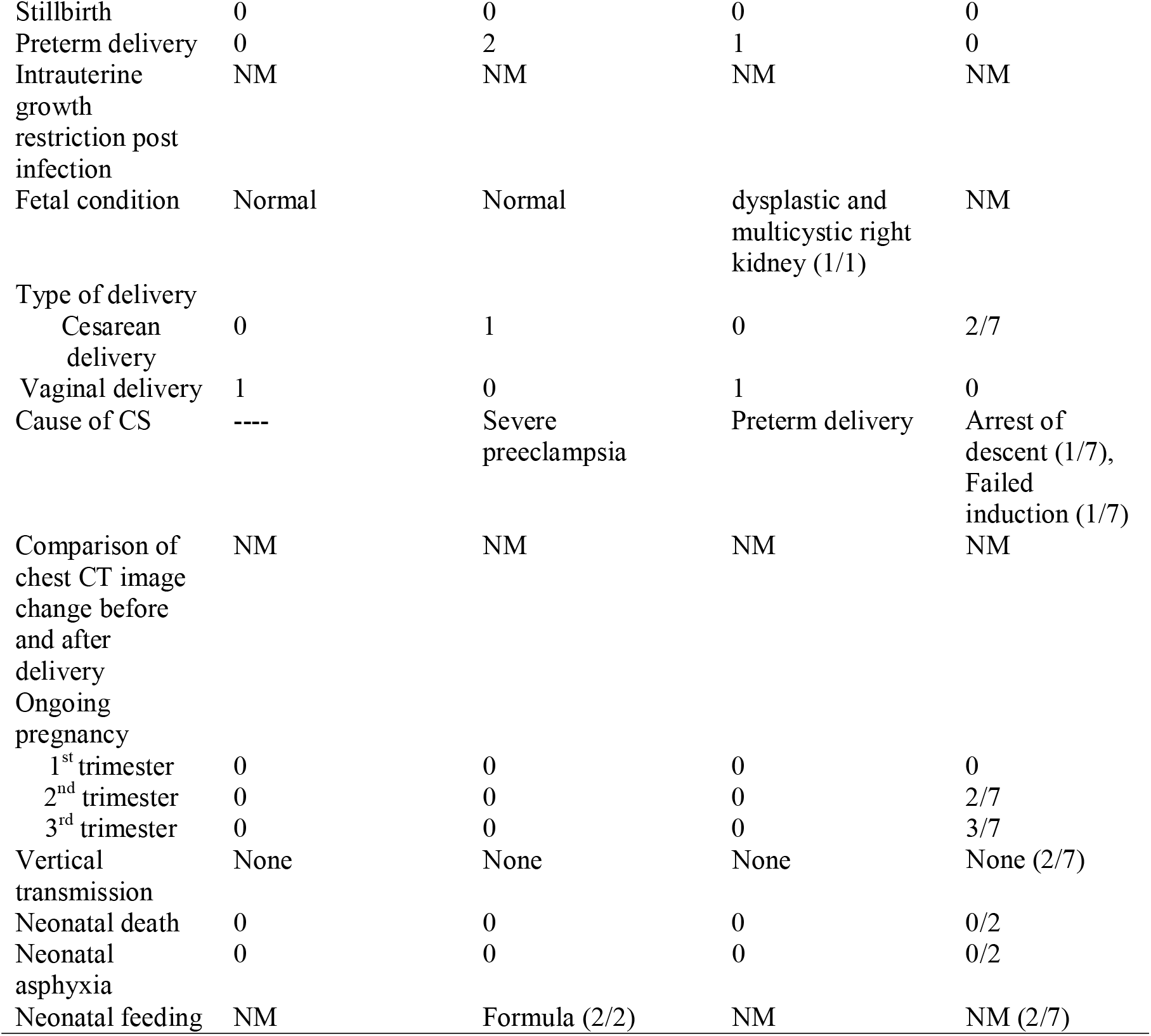
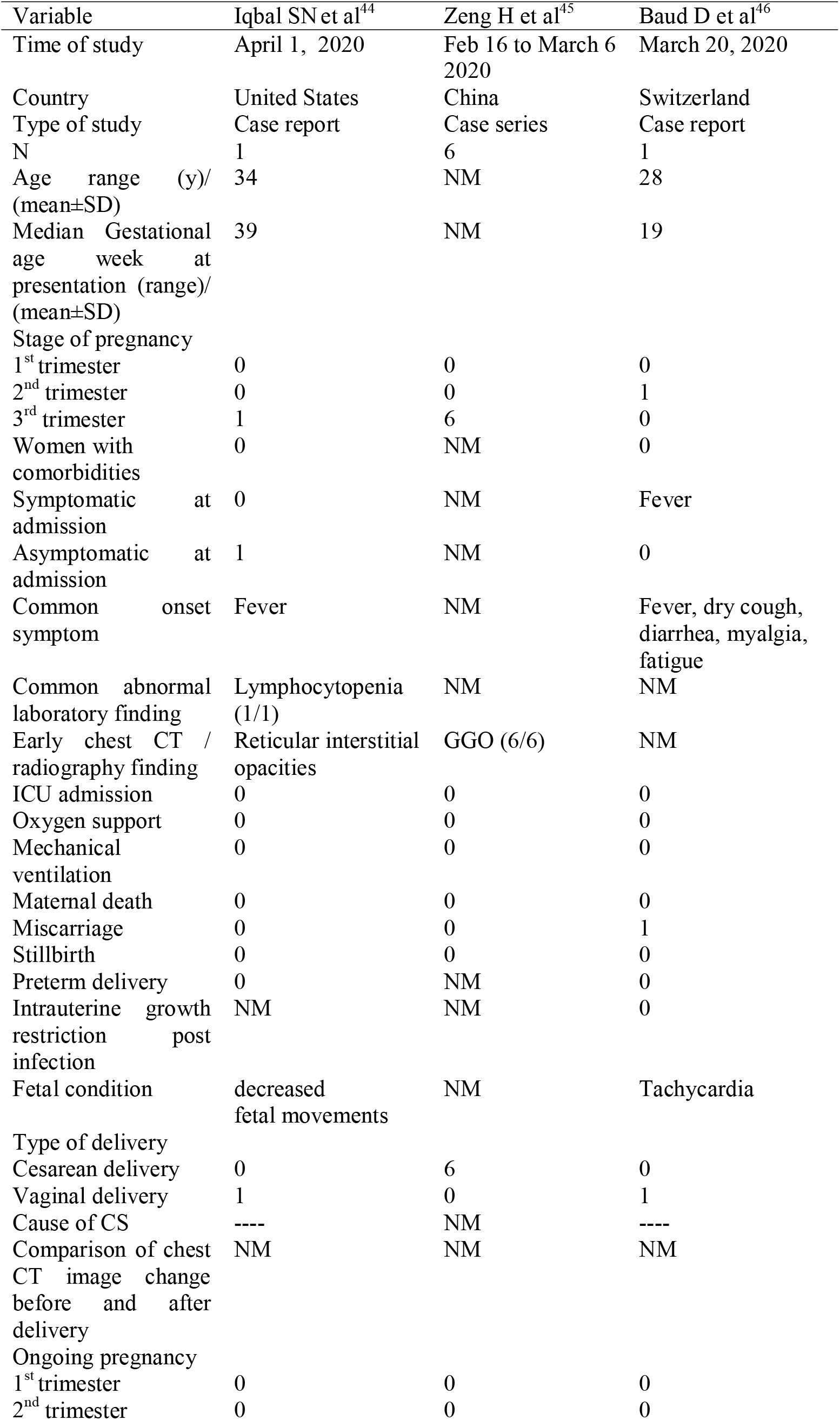
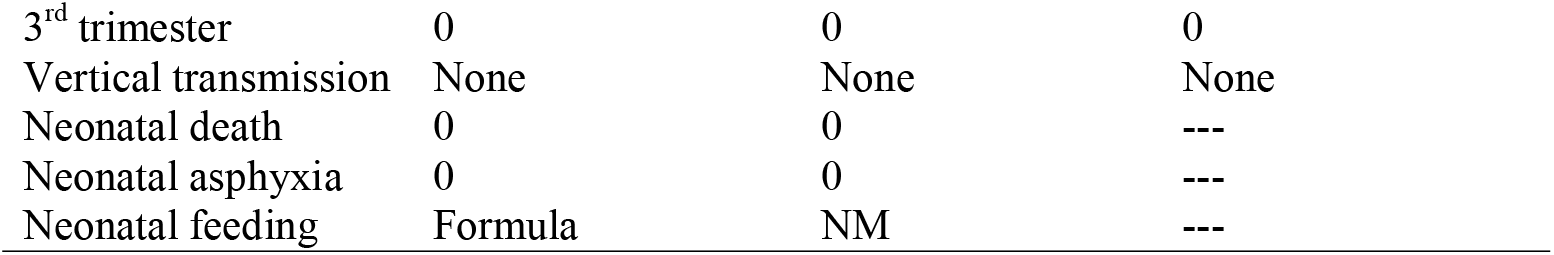
Characteristics of studies included in COVID-19 impact review on maternal and neonatal outcomes

## 3. Results

A flow diagram of the literature search is shown in Figure 1. From 77 results in PubMed, 13 in Scopus, 7 in WOS, and 45 in MedRxiv, 28 relevant studies (15 case series, 10 case reports, 1 case-control, 2 Retrospective clinical series) were identified. Characteristics of the selected studies are described in Table 1. Two full texts of included articles are in Chinese [26,38] and the others are in English. All articles reported SARS-CoV-2 positive pregnant women from China except six case reports from Republic of Korea (1) [39], Sweden (1) [41], United States (2) [44,45], Central America (1) [42], and Switzerland (1) [46]. A total of 287 pregnant women with COVID-19 were identified.

**Figure 1:**
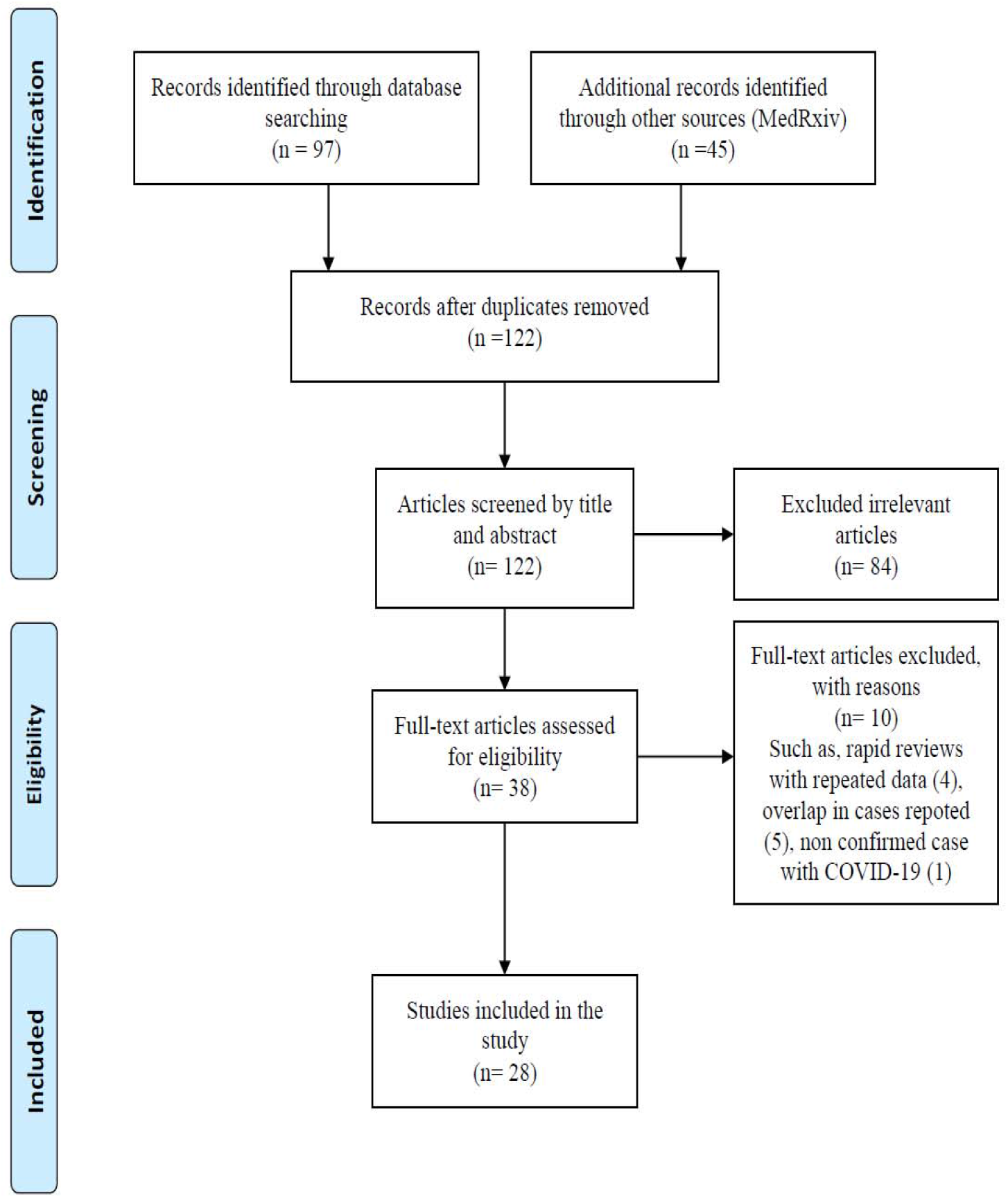
Flow diagram of literature search.

### Maternal characteristics

Mean maternal age ranged from 22 to 41 years of age. Most pregnant women reported in their third trimester. Eleven pregnant women (3.8 %) were in their second trimester and there was no data on first-trimester COVID-19 infection. All pregnancies were singleton except 5 twin cases. Forty-two pregnancies (15%) were ongoing including seven women (2.5%) in second trimester, ten women (3.6%) in third trimester, and trimester of twenty-five women (8.9%) was not mentioned. All these women were discharged without any major complications. The outcomes of these pregnancies are not known.

Sixty-seven pregnant women (23.9%) had co-morbidities or complications in their pregnancies. Gestational diabetes mellitus, gestational hypertension, and preeclampsia were the most common complications in these pregnancies. At the time of admission, 102 (35.5%) and 34 (11.8%) cases were symptomatic and asymptomatic, respectively. The status of symptoms at the time of admission was not mentioned in 152 of 287 cases. Fever was a common onset symptom in pregnant women with COVID-19 (51.5%), although onset symptoms were not mentioned in 22 cases. Lymphocytopenia was a common abnormal laboratory finding which was reported in 91 of 134 cases (67.9%) where the information was recorded. Multiple ground-glass opacities were the most common pattern visible in chest computed tomography (CT) (201 of 256 available cases).

### Maternal outcomes

There is currently no data about first trimester miscarriage affected women with COVID-19 infection because all pregnant women were in second and third trimester of their pregnancies. In terms of second or third-trimester pregnancy loss, there was only one stillbirth in a woman at 34 weeks gestational age with a fever and sore throat whose condition deteriorated during hospitalization, she was transferred to ICU and required Extracorporeal Membrane Oxygenation (ECMO) due to multiple organ dysfunction syndrome (MODS) [27]. In addition, a second-trimester miscarriage in a pregnant woman with COVID-19 infection was reported at 19 weeks gestational age [46]. Forty-three women delivered preterm. 88.38% of these preterm deliveries were delivered electively by cesarean section. Fetal intrauterine growth restriction post COVID-19 infection was not reported in any studies because pregnant women delivered a short time after onset of illness and no data were available regarding to fetal growth monitoring in ongoing pregnancies at the time of publication. Placental pathology in three cases showed that lack of morphological changes was related to viral infection, no villitis or chorioamnionitis, and negative 2019-nCoV nucleic acid test [38]. Although, positive RT-PCR test for SARS-Cov-2 of placental submembrane and cotyledon was reported in a pregnant woman with COVID-19 in second trimester of her pregnancy [46]. 93% of all deliveries were done through cesarean section and vaginal delivery was reported in 17 of 243 cases. According to the mentioned causes of cesarean section in 84 cases, COVID-19 infection was the most common indication for cesarean section in 44 of 84 cases (52.38%), and fetal distress was ranked second with 13 of 84 cases (15.47%). From the papers that provided information about fetal condition assessment (118 cases), fetal distress was reported in 20 cases (16.9%) and fetal condition in 94 cases (79.6%) was reported normal. From all pregnant women with COVID-19, 156 cases received oxygen support through a nasal cannula and mask and 5 cases were admitted to the intensive care unit (ICU) and 4 women required intubation and mechanical ventilation. No maternal death was reported to date.

### Neonatal outcomes

Neonatal asphyxia was reported in 7 of 232 cases (3%). A male newborn death was reported on the 9th day after birth [30]. His mother developed thrombocytopenia complicated with abnormal liver function. He born at 34 weeks and 5 days gestational age with an Apgar score of 8 at five minutes and admitted to NICU due to shortness of breath and moaning. On the 8th day after birth, his condition deteriorated due to refractory shock, multiple organ failure, and disseminated intravascular coagulation (DIC). Result of his throat swab for 2019-nCoV nucleic acid testing was negative on 9th day after delivery. According to the mentioned newborn feeding in 60 cases, all newborns of mothers with COVID-19 were given formula instead of breast milk. Vertical transmission of COVID-19 from mother to the fetus was not reported in available literature. However, possibility of mother-to-child vertical transmission was suggested in three cases [23,29,35]. The first one was a newborn with elevated IgM level, cytokines, and white blood cell count, normal chest CT, and negative RT-PCR tests on nasopharyngeal swabs taken from 2 h to 16 days of age [23]. The second one was a neonate with positive nucleic acid test of SARS-CoV-2 at 36 h after birth, mild shortness of breath symptoms, mild pulmonary infection in chest X-ray and no fever and cough. The neonate was discharged after 2 weeks following two consecutive negative nucleic acid test results [29]. The third one was a neonate with positive RT-PCR test for SARS-Cov-2, pulmonary infection in chest X-ray 53 h after birth, and no clinical manifestations of Covid-19. The neonate was discharged after subsequent negative RT-PCR of throat swabs on 16^th^ day after birth [35]. Additionally, two newborns were reported with elevated SARS-CoV-2 IgM antibodies, negative RT-PCR test results of neonatal throat swabs, and no Covid-19 infection symptoms [45].

## 4. Discussion

After the first COVID-19 pneumonia cases were reported in Wuhan, Hubei Province, China, in December 2019, and the ongoing outbreak across the world [1], some reports in pregnant women with COVID-19 infection have been identified. This article summarizes available information from 287 pregnant cases with COVID-19.

Studies showed that SARS infection was associated with 1^st^ trimester spontaneous abortion, fetal growth restriction due to fibrin deposition, and a high case fatality rate (CFR) of 25% in pregnant women [47,48]. A case-control study done in Hong Kong reported that ICU admission rate and CFR in the pregnant group were higher than non-pregnant group and were 60% and 40% respectively [49]. Limited data exists on the outcome of MERS on pregnancy, however, available information showed that CFR of infected pregnant women was 27% and ICU admission was 64% [4, 50-54]. Our review of 287 pregnant women with COVID-19 showed no maternal mortality and ICU admission was reported in 5 of 161 cases (3%).

Studies showed that the decision for cesarean section in SARS and MERS affected pregnant women was made due to maternal hypoxemia, fetal distress, and some obstetrical indications such as placenta previa [4,51,55,56]. In this review, the majority of deliveries were done through cesarean section and 6.9% of pregnant women delivered by vaginal delivery. More than half of mentioned causes of cesarean section were COVID-19 infection and 15.4% of deliveries were done through cesarean section due to fetal distress. One important issue was that in most women who delivered by cesarean section due to COVID-19 infection cause, maternal condition did not deteriorate.

Vertical transmission from mother to child among pregnant women who were infected with SARS and MERS were not identified [4,57-60]. According to this review, there is no evidence to prove intrauterine or transplacental transmission of COVID-19 but possibility of mother-to-child vertical transmission was suggested due to positive RT-PCR test for SARS-Cov-2 in two newborns [29,35], elevated SARS-Cov-2 IgM anitibodies in three neonates alongside negative RT-PCR [23,45], and virological findings in placenta in one case [46]. But some researchers stated that molecular tests based on nucleic acid amplification and detection are more reliable than IgM test to diagnose infections due to false-positive and false negative results, along with cross-reactivity and testing challenges of IgM assays [61].

Although no viral RNA was detected in breast milk of mothers infected with SARS and MERS who had been tested by RT-PCR, breast feeding in these mothers was not advised [4,53,56]. Chinese expert consensus on the perinatal and neonatal management for the prevention and control of Covid-19 infection states that mothers with confirmed or who are suspected of Covid-19 should not feed their infants with breast milk. However, if the suspected or diagnosed mother and her breast milk test is negative for Covid-19, infants should be fed with breast milk [62]. Though COVID-19 has not been detected in breast milk samples in some studies [20,22,23,25,31], all newborns of mothers with COVID-19 were not breastfed according to available information. It seems that formula feeding could prevent neonates from the possible way of Covid-19 infection transmission through close contact with confirmed or suspected mothers.

Lack of report of some variables related to maternal and neonatal outcomes in several studies is one of the limitations of this review. The other one is low methodological quality of the studies which were included in this review. Maternal and neonatal assessment of a large number of cases (287 cases), consideration of some maternal and neonatal outcomes including cause of cesarean section, early maternal chest CT findings, fetal condition, neonatal asphyxia, newborn feeding in all studies and no limitation on language of published articles are strengths of the current review in comparison with three reviews that exist to date [63-66].

## Conclusion

This review literature showed fewer adverse maternal and neonatal outcomes in pregnant women with COVID-19 in comparison with SARS and MERS infection in pregnancy. There is no evidence to prove vertical transmission of COVID-19 but possibility of mother-to-child vertical transmission was suggested due to positive test for SARS-Cov-2 in two neonates and one placental infection with SARS-Cov-2. Possibility of transmission during vaginal delivery is unknown due to small number of cases who delivered by vaginal delivery. Because of not reporting breastfeeding, risk of transmission during breast feeding is unknown if breast milk test of suspected or diagnosed mother with Covid-19 is negative.

## Data Availability

All data are available in the text of manuscript.

## Declaration of Competing Interest

The authors of this study declare that they each have no conflict of interest

## Funding Statement

This research did not receive any specific grant from funding agencies in the public, commercial, or not-for-profit sectors.

## Appendix 1

### PubMed search strategy

(“2019-nCoV infection”[all] OR “coronavirus disease 2019”[all] OR “COVID-19 pandemic”[all] OR “2019-nCoV disease”[all] OR “2019 novel coronavirus disease”[all] OR “COVID19”[all] OR “2019 novel coronavirus infection”[all] OR “coronavirus disease-19”[all] OR “severe acute respiratory syndrome coronavirus 2”[all] OR “SARS-CoV-2”[all]) AND (pregnancy[all] OR “pregnant women”[all] OR maternal[all] OR “prenatal care”[all]) AND (2019/10/01: 2020/04/30 [dp])

### Scopus search strategy

(ALL(“2019-nCoV infection”) OR ALL(“coronavirus disease 2019”) OR ALL(“COVID-19 pandemic”) OR ALL(“2019-nCoV disease”) OR ALL(“2019 novel coronavirus disease”) OR ALL(“COVID19”) OR ALL(“2019 novel coronavirus infection”) OR ALL(“coronavirus disease-19”) OR ALL(“severe acute respiratory syndrome coronavirus 2”) OR ALL(“SARS-CoV-2”)) AND (ALL(pregnancy) OR ALL(“pregnant women”) OR ALL(maternal) OR ALL(“prenatal care”)) AND (PUBDATETXT(“October 2019” OR “November 2019” OR “December 2019” OR “January 2020” OR “February 2020” OR “March 2020” OR “April 2020”))

### WOS search strategy

(ALL=(“2019-nCoV infection”) OR ALL=(“coronavirus disease 2019”) OR ALL=(“COVID-19 pandemic”) OR ALL=(“2019-nCoV disease”) OR ALL=(“2019 novel coronavirus disease”) OR ALL=(“COVID19”) OR ALL=(“2019 novel coronavirus infection”) OR ALL=(“coronavirus disease-19”) OR ALL=(“severe acute respiratory syndrome coronavirus 2”) OR ALL=(“SARS-CoV-2”)) AND (ALL=(pregnancy) OR ALL=(“pregnant women”) OR ALL=(maternal) OR ALL=(“prenatal care”)) AND PY=(2019-2020)

## Notes

### Competing Interest Statement

The authors have declared no competing interest.

### Funding Statement

This work was supported by the Vice Chancellor for Research, Shiraz University of Medical Sciences, Shiraz, Iran.

